# Computed tomography analysis of vulnerable carotid atherosclerotic plaque and relationship to clinical characteristics

**DOI:** 10.1101/2022.11.01.22281634

**Authors:** N. Giannotti, J.P. McNulty, S.J. Foley, M. Barry, M. Crowe, E. Dolan, J. Harbison, G. Horgan, E. Kavanagh, M O’Connell, M. Marnane, S. Murphy, C. McDonnell, M. O’Donohoe, D. Williams, P.J. Kelly

## Abstract

**Objective:** The presence of atherosclerotic plaque components such as lipid rich necrotic core and intraplaque haemorrhage is associated with increased plaque vulnerability, and may be used to stratify the risk of future cerebrovascular events. Our aim was to investigate the relationship between selected carotid plaque components imaged with CTA, patient characteristics, and clinical outcomes.

**Methods:** Symptomatic patients underwent carotid CTA as part of the BIOVASC study. Images were analysed for plaque volume composition with a semi-automatic Hounsfield Unit (HU)-based algorithm. Plaque components were classified based on their attenuation values: lipids <61 HU, fibrous tissue 61-129 HU and calcium >131 HU. Parametric and non-parametric tests were performed to compare plaque measurements to clinical characteristics and outcomes.

**Results:** One-hundred and two symptomatic carotids were analysed (avg. age 69y, 54.9% Male, 29.4% severe stenosis). Mean plaque volume was 480±230 mm^3^, and the mean LRNC volume was 170±100 mm^3^. A difference in LRNC volume was identified between moderate and severe stenosis (190–150 mm^3^, p=0.012). Regression analysis showed that age and gender may predict increased plaque volume (p<0.001). A trend for reduced mean plaque LRNC was identified in patients receiving statins (130-210 mm^3^, p=0.08). Intra-reader reliability showed good agreement (0.62-0.78, p<0.001) between CTA plaque measurements.

**Conclusions:** In-vivo CTA plaque volume composition assessment is feasible with good intra-reader reliability. Our findings suggest that CTA-HU measurements may be used to provide improved mechanistic and diagnostic insights into atherosclerotic disease, and facilitate the quantification of selected plaque components whose presence may be associated with increased plaque vulnerability.

**Key Points:**

- Plaque CTA Hounsfield Unit (HU) measurement and segmentation techniques can provide improved mechanistic and diagnostic insights into atherosclerotic disease.
- Plaque components characterisation using CTA HU measurements may help clinicians to identify patients presenting with high-risk vulnerable plaque.
- CTA plaque volume composition assessment is feasible with good reliability observed between measurements taken at different time-points.

## Introduction

The World Health Organization (WHO) identified cardiovascular diseases as the leading cause of death globally [1]. Cardiovascular diseases account for most noncommunicable diseases (NCDs) deaths, or 17.9 million people annually, followed by cancers (9.3 million), respiratory diseases (4.1 million), and diabetes (1.5 million) [1]. Stroke alone accounts for an estimated 6.2 million and was ranked as the second leading cause of global death [1]. Large, randomised trials including the North American Symptomatic Carotid Endarterectomy Trial (NASCET) and the European Carotid Surgery Trial (ECST) demonstrated that carotid revascularisation reduces stroke risk in patients with symptomatic severe carotid stenosis (> 70%) [2,3], but has marginal impact in several sub-groups of patients with moderate and mild stenosis [4].

Today, carotid luminal stenosis - measured with digital subtraction angiography (DSA), computed tomography (CT), magnetic resonance imaging (MRI), and ultrasound (US) - represents the primary determinant of treatment for patients with carotid artery stenosis. However, histopathologic studies showed that carotid plaques with identical luminal stenosis were found to be histologically different suggesting that plaque composition may trigger the increased risk of ischaemic event [5,6]. For this reason, vessel wall imaging has been increasingly used to assess plaque beyond luminal stenosis to identify those patients who may be at higher risk of cerebrovascular ischemia [7,8,9].

In the settings of ischaemic stroke, CT Angiography (CTA) is indicated for carotid imaging due to its wide availability, relative low-cost and excellent inter-operator agreement [10]. In addition to accurately assessing carotid plaque wall and luminal stenosis, CTA has been recently used to investigate selected plaque biomarkers that may be associated with plaque vulnerability and that may support patient management decisions [11,12]. Using a dataset from the prospective Biomarkers Imaging Vulnerable Atherosclerosis in Symptomatic Carotid disease (BIOVASC) study [13], this study investigated the feasibility of CTA to segment and quantify selected carotid plaque biomarkers and explored their relationship with clinical characteristics and outcomes.

## Materials and methods

### Eligibility criteria

Pre-specified inclusion criteria were: (1) age ≥50 years; (2) presentation to medical attention with recent (<30 days) non-severe ischaemic stroke (modified Rankin scale [MRS] ≤3) or motor/speech/vision TIA; (3) ipsilateral internal carotid artery (ICA) stenosis (≥ 50% lumen-narrowing) on admission doppler US, MRA or CTA done for clinical care; (4) CTA completed. Main exclusion criteria were: (1) possible haemodynamic stroke/TIA due to carotid near-occlusion; (2) contraindication to contrast-enhanced CT (serum creatinine level > 1.5 mg/dL or estimated glomerular filtration rate < 60mls/min) [13]. All symptomatic patients were clinically examined and underwent carotid CTA at the date of medical presentation; risk factors and baseline characteristic were collected (Table 1). The study was approved by the Institutional Review Board with written informed, consent for all patients.

**Table 1.**
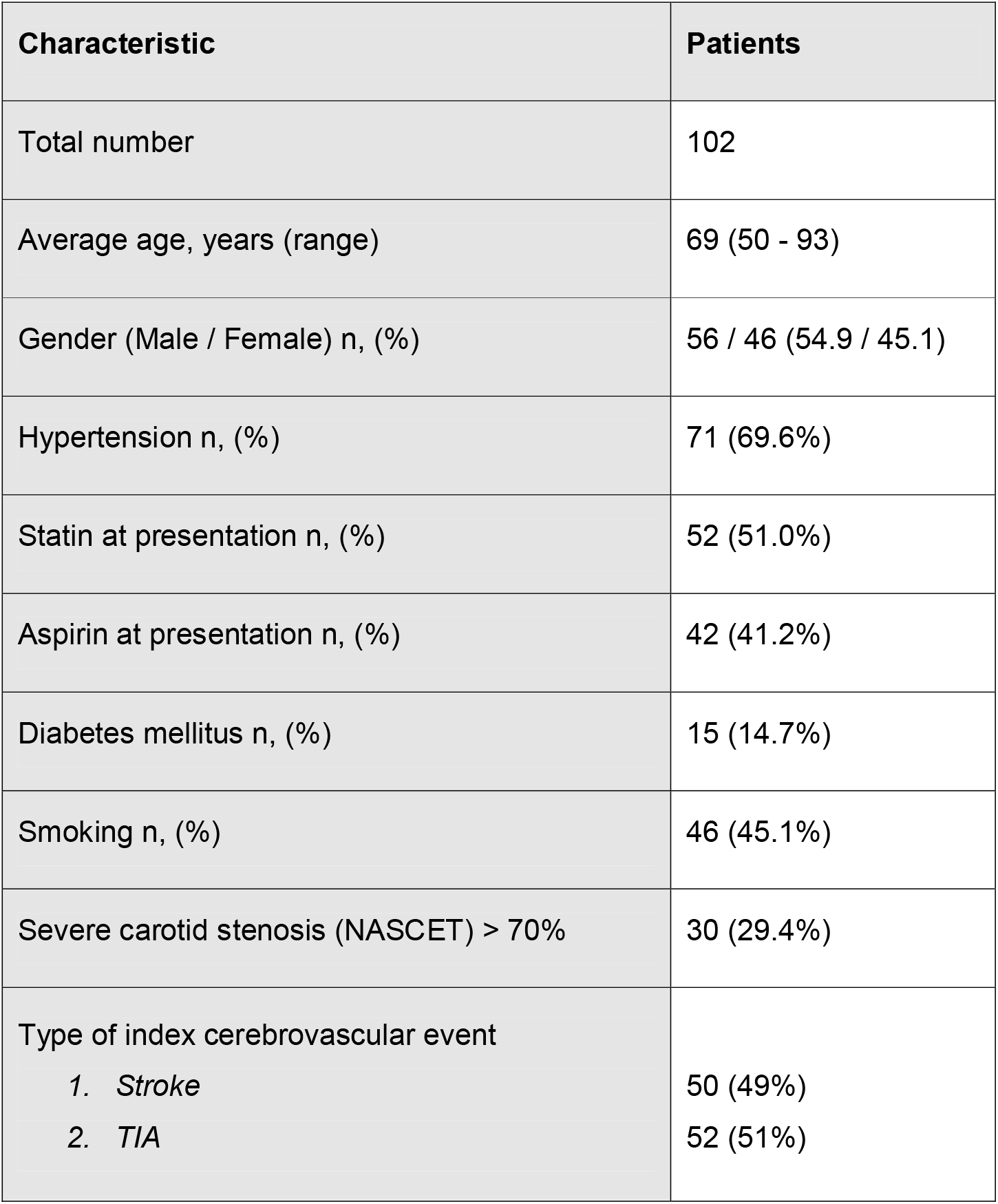
Clinical baseline characteristics of patients.

| Characteristic | Patients |
| --- | --- |
| Total number | 102 |
| Average age, years (range) | 69 (50 - 93) |
| Gender (Male / Female) n, (%) | 56 / 46 (54.9 / 45.1) |
| Hypertension n, (%) | 71 (69.6%) |
| Statin at presentation n, (%) | 52 (51.0%) |
| Aspirin at presentation n, (%) | 42 (41.2%) |
| Diabetes mellitus n, (%) | 15 (14.7%) |
| Smoking n, (%) | 46 (45.1%) |
| Severe carotid stenosis (NASCET) $> 70\%$ | 30 (29.4%) |
| Type of index cerebrovascular event |  |
| 1. *Stroke* | 50 (49%) |
| 2. *TIA* | 52 (51%) |

### CTA protocol

CTA data were collected from six participating centres using Siemens Biograph (Siemens Healthineers, Erlangen, Germany), GE Healthcare System Discovery 690 (Milwaukee, Wisconsin) and Philips Healthcare Gemini (Philips, Eindhoven Netherland). All scanners were multi-detectors (16 – 64 slices).

CTA carotid acquisition protocols were standardised to ensure accuracy and consistency in data collection and analysis. Ninety millilitres of Omnipaque 350 (GE Healthcare Systems) were injected at a flow rate of 4 mL/s in the antecubital vein, subsequently a diagnostic carotid CTA was performed using bolus tracking. CT images (0.66-0.75 mm) with contrast-enhancement were acquired from the aortic arch to the skull base to identify carotid arteries and jugular veins. CTA images were reconstructed into 1 mm slices and the smooth– medium reconstruction kernel was used across the scanning centres: b30f (Siemens Healthineers Biograph), C (Philips Healthcare Gemini) and STANDARD (GE Healthcare Systems Discovery 690).

### Image analysis

CTA images were analysed by a single reader with 3 years’ experience in carotid imaging. Curved-planar reformation (CPR) images were used to navigate along the carotid artery, and the degree of each symptomatic carotid stenosis was confirmed on the correspondent CTA axial section showing the maximal luminal narrowing (Fig. 1). As per NASCET criteria, measurements of distal ICA diameters were taken beyond the bulb where the wall is parallel [2,14]. A window width and level of 800 / 300 HU respectively were used to differentiate the contrast filled lumen from the carotid vessel wall [14].

**Figure 1.**
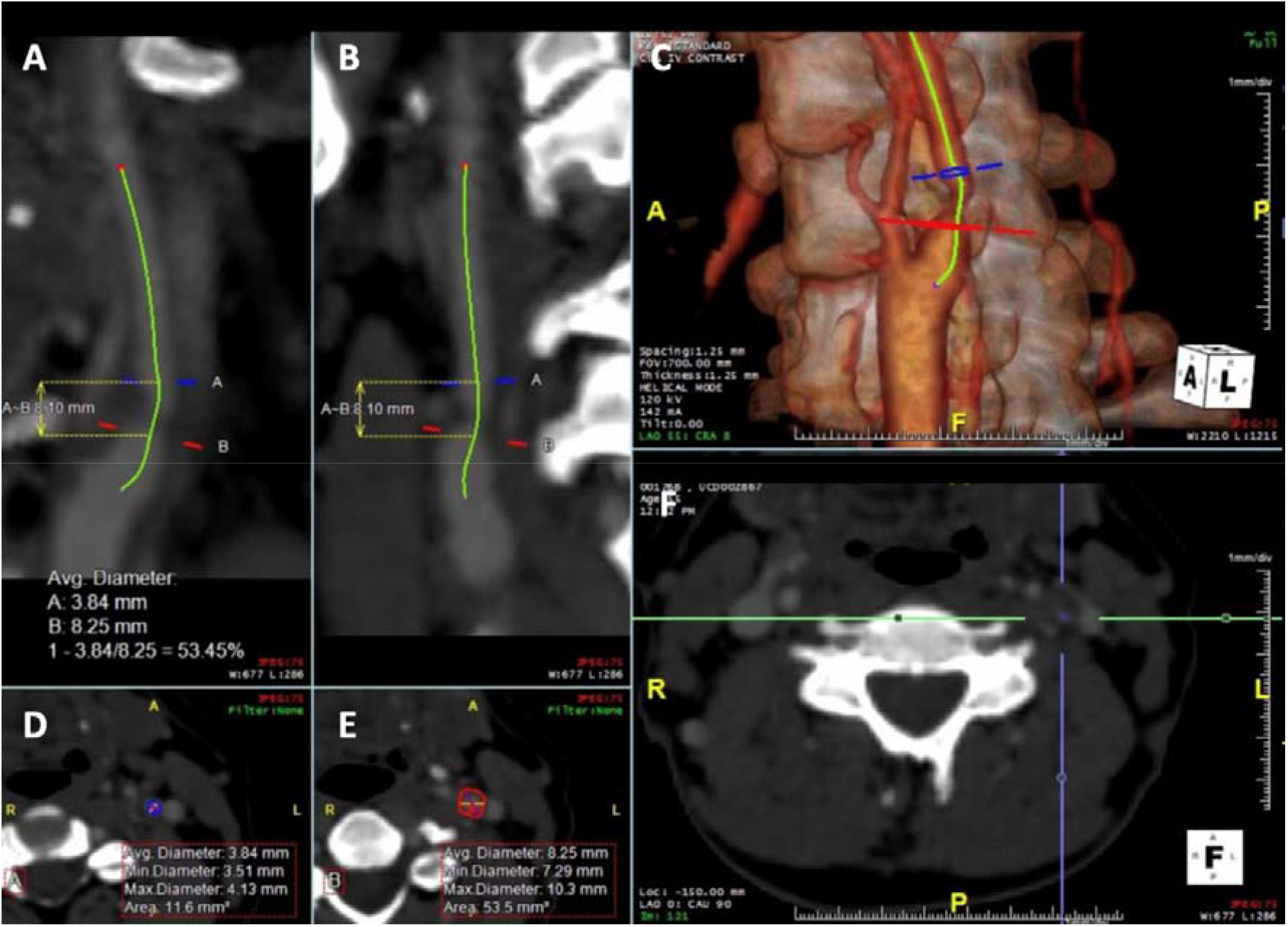
Semi-automated analysis of left internal carotid artery (ICA) stenosis using CT angiography. A and B: Curved planar reformatted images (CPR) taken at different angles of the left ICA showing the length of the stenosis - proximal (blue) and distal (red) stenosis boundaries. C: left ICA 3D reconstruction. D and E: axial sections of the proximal (blue) and distal (red) boundaries of the ICA stenosis. F: CTA axial slice showing the point of maximum ICA stenosis.

Carotid atherosclerotic plaque was defined as the area between inner and outer boundaries of a thickened vessel. The software package *“Plaque analysis”* available with AquariusNet iNtuition™ (TeraRecon) was used for the semi-automatic HU-based analysis of the plaque components. Distal and proximal boundaries were drawn manually along the vessel to enable the software to perform a semi-automatic segmentation of the vessel wall from the lumen along the selected ICA volume. Manual adjustments were made on the axial plane when necessary. Consequently, three histologically validated HU ranges [15,16] were applied to obtain volumetric measurements of plaque components within the vessel wall as follows: < 61 HU for lipid, 61 to 130 HU for fibrous tissue, HU > 131 for calcium (Fig. 2).

**Figure 2.**
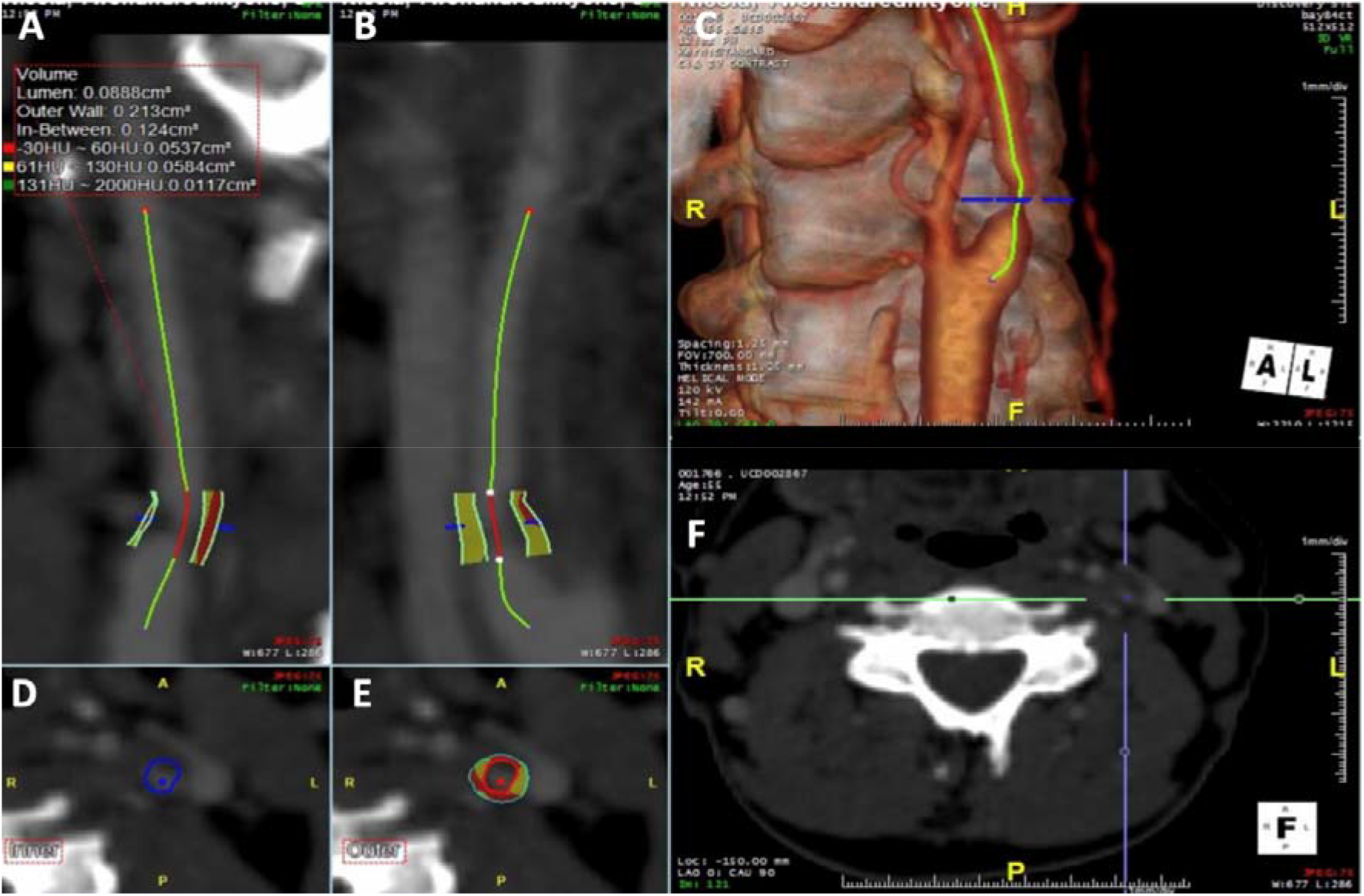
Curved planar reformatted CTA showing a severe left ICA stenosis. A and B: pre-defined HU range colour map applied to the ICA stenosis. C: ICA 3D reconstruction. D: luminal boundaries. E: plaque wall boundaries with HU range colour map. F: CTA axial slice showing the point of maximum ICA stenosis. Plaque view was able to identify and quantify the different plaque components: LRNC (yellow), fibrous tissue (red) and calcium (green).

### Statistical analysis

Normality of data was assessed with Shapiro-Wilk test. The intraclass correlation (ICC) test was used to assess the intra-reader reliability by repeating the analysis on 25% of the total dataset following a six-month interval. Parametric and non-parametric tests were performed based on the distribution of data. All statistical analyses were performed with SPSS Version 21 (IBM Corp, NY, USA).

## Results

One-hundred and two symptomatic carotids were analysed. Atherosclerotic lesions were categorised according to NASCET criteria into four groups: mild (< 50%), moderate (50% – 69%), severe (70% - 95%) and near-occlusion (> 95%). Across the four stenosis categories, the mean volume for lipid was 170 mm^3^, for fibrous tissue was 140 mm^3^ and for calcification was 170 mm^3^ (Table 2).

**Table 2.** Volumetric distribution (%) of plaque components among stenosis categories.

| **Stenosis**<br>**NASCET** | **N (%)** | **Mean**<br>**plaque vol**<br>**(mm<sup>3</sup>) ±SD** | **Mean lipid**<br>**vol (mm<sup>3</sup>)**<br>**±SD** | **Mean fibrous**<br>**tissue vol**<br>**(mm<sup>3</sup>) ±SD** | **Mean**<br>**calcium vol**<br>**(mm<sup>3</sup>) ±SD** |
| --- | --- | --- | --- | --- | --- |
| **Mild (< 50%)** | 8 (7.5) | 310 ± 170 | 130 ± 80 | 100 ± 60 | 90 ± 30 |
| **Moderate**<br>**(50% – 69%)** | 43 (42.5) | 470 ± 90 | 190 ± 110 | 150 ± 80 | 150 ± 50 |
| **Severe**<br>**(70% - 95%)** | 42 (41.5) | 580 ± 140 | 150 ± 7 | 130 ± 70 | 180 ± 80 |
| **Near-occlusion**<br>**(> 95%)** | 9 (8.5) | 710 ± 210 | 270 ± 170 | 150 ± 100 | 290 ± 120 |
| **Whole group** | 102 (100) | 480 ± 230 | 170 ± 100 | 140 ± 100 | 170 ± 120 |

The Mann-Whitney U test showed greater LRNC volume in moderate stenosis compared to severe stenosis (190 mm^3^-150 mm^3^, p=0.012). Kruskal-Wallis test did not show any significant variation between volumetric measurements of plaque fibrous tissue (p=0.44) and calcifications (p=0.25) across the different categories of stenosis. After adjusting for confounders such as smoking, diabetes, therapeutic treatments and degree of stenosis, multiple regression analyses were calculated to predict plaque volume based on age and gender. Results of the regression analysis showed that these variables may predict increased plaque volume F = 7.74, p<0.001 and R^2^ = 0.37, and they both added statistically significantly to the prediction (p<0.05). Plaque mean volume was larger in males compared to females (550 mm^3^ vs 300 mm^3^, p< 0.001); moreover, a significantly larger volume of plaque LRNC was found among male patients when compared to the female group (210 mm^3^ vs 100 mm^3^, p < 0.001). A trend for reduced mean plaque LRNC was identified in patients receiving statins (130-210 mm^3^, p=0.08) (Table 3). The intra-reader reliability, measured with the ICC test, showed overall good overall agreement for all the HU ranges and plaque features analysed as shown in Table 4.

**Table 3.** Relationships between selected plaque morphological features and patient characteristics (102 patients).

|  | **Plaque**<br>**volume**<br>**(mm<sup>3</sup> ± SD)** | **p** | **LRNC (mm<sup>3</sup>**<br>**± SD)** | **p** | **Calcium**<br>**(mm<sup>3</sup> ± SD)** | **p** |
| --- | --- | --- | --- | --- | --- | --- |
| **Hypertension** |  |  |  |  |  |  |
| Yes | 520 ± 400 | .07 | 170 ± 130 | .81 | 200 ± 120 | **0.01** |
| No | 390 ± 230 |  | 170 ± 140 |  | 80 ± 60 |  |
| **Statin** |  |  |  |  |  |  |
| Yes | 470 ± 390 | .80 | 130 ± 87 | .08 | 200 ± 124 | .054 |
| No | 490 ± 350 |  | 210 ± 120 |  | 120 ± 86 |  |
| **Gender** |  |  |  |  |  |  |
| Male | 550 ± 390 | **.001** | 210 ± 78 | **.001** | 180 ± 140 | .07 |
| Female | 300 ± 140 |  | 100 ± 75 |  | 120 ± 100 |  |
| **Diabetes** |  |  |  |  |  |  |
| Yes | 430 ± 200 | .60 | 150 ± 120 | .30 | 140 ± 80 | .62 |
| No | 490 ± 390 |  | 180 ± 130 |  | 170 ± 84 |  |
| **Smoking** |  |  |  |  |  |  |
| Yes | 490 ± 390 | .32 | 180 ± 113 | .47 | 170 ± 122 | .20 |
| No | 390 ± 200 |  | 150 ± 111 |  | 100 ± 60 |  |
| **Type of event** |  |  |  |  |  |  |
| Stroke | 540 ± 440 | .12 | 200 ± 112 | .12 | 190 ± 126 | .22 |
| TIA | 420 ± 290 |  | 140 ± 120 |  | 140 ± 89 |  |

**Table 4.** ICC test of three different plaque components (measurements were taken after 6-month time interval from the initial analysis).

| HU ranges | Mean mm<sup>3</sup> ± SD | ICC (α) | 95% CI min | 95% CI max | p |
| --- | --- | --- | --- | --- | --- |
| **Lipid (-30 / 60) HU** | 180 ± 70 | 0.65 | 0.54 | 0.71 | < 0.001 |
| **Fibrous tissue (61 / 130) HU** | 140 ± 60 | 0.62 | 0.50 | 0.77 | < 0.001 |
| **Calcium (> 130) HU** | 170 ± 100 | 0.78 | 0.65 | 0.86 | < 0.001 |
| **Plaque volume** | 1571 ± 526.4 | 0.72 | 0.79 | 0.79 | < 0.001 |

## Discussion

In agreement with current literature, this study extends the knowledge supporting CTA as a reliable diagnostic tool capable of quantifying selected plaque components that have histologically been associated with plaque vulnerability [17]. *In-vivo* evaluation of carotid plaque beyond luminal stenosis is becoming a well-known technique able to provide a comprehensive assessment of plaque morphology and may become helpful in patient risk-stratification.

Carotid plaque volume in this study was significantly larger in males than females; moreover, carotid plaque in the male group seemed to be more complex compared to the female group due to their larger volume of LRNC. Although further research is needed, these findings support the evidence that in women hormones may have a key protective role in plaque stabilisation [18]. This study also identified age as an independent predictor of plaque volume increase, which correlates to the faster carotid plaque progression rate occurring in the older population [19].

In this analysis we found larger volumes of LRNC in the cohort of symptomatic patients presenting with moderate stenosis and a trend for reduced mean plaque LRNC volume was identified in patients receiving statins. The most immediate implication of such findings is that despite causing a moderate vessel occlusion, these plaques should be considered at a higher risk of causing ischaemic events such as stroke or TIA. Nevertheless, plaque biomarkers are not commonly used in clinical settings where the degree of carotid artery stenosis is still the most common criteria used to predict the risk of cerebrovascular events in patients affected by carotid atherosclerosis.

In this study we did not cross-validate the CTA findings with histological specimens, and we adopted pre-defined plaque-HU ranges that previously showed good association with corresponding plaque histology [6,16]. CTA has always been considered an accurate diagnostic test for the identification of several carotid morphological features such as luminal narrowing, intraplaque calcifications and ulcerations [6]; however, CTA plaque components characterisation mostly remains a research tool with limited application in clinical practice [20,21]. The HU values that we used for plaque soft component characterisation (<61 HU for lipid) did not discriminate attenuation differences between intra-plaque haemorrhage (IPH) and LRNC; therefore, this study limited the CTA carotid plaque analysis to the identification of only three plaque components: LRNC, fibrous tissue and calcium. The detection with CTA of other plaque components associated with plaque vulnerability such as IPH and fibrous cap (FC) is challenging and conflicting results were reported [6,22]. These plaque components have been associated with increased risk of cerebrovascular ischaemic event [23]; however, due to spatial resolution limitations and similar attenuation coefficients CTA has not been validated yet as a LRNC – IPH characterisation tool in clinical practice. The current literature acknowledges MRI as the imaging modality of choice in the detection of LRNC [6,7]; however, low availability, high cost and long scanning time are some of the limitations of carotid MRI in acute stroke settings. For these reasons we believe that further studies on large cohorts of patients should be supported to confirm the potential of CTA as a plaque characterisation tool.

The recent introduction of Photon-Counting Computed Tomography (PC-CT) scanners in clinical practice may have the potential to significantly change clinical CTA and plaque imaging. Briefly, PC-CT relies on detectors that do not require a separate step to convert the X-rays into light [24]. Each photon that hits the detector of a PC-CT system produces an electrical pulse with a height proportional to the energy deposited by the photon; therefore, PC-CT can count the number of incoming photons and measure their individual energy [24]. This technique results in improved spatial resolution, higher contrast-to-noise ratio, correct beam-hardening artifacts, lower radiation exposure and optimized spectral imaging [25]. For these reasons PC-CT may have the potential to overcome most of the current technical demands of plaque imaging associated with high-resolution requirements and image noise. We believe that further research in the settings of non-invasive high-risk plaque imaging should be performed to explore whether PC-CT may have the potential to characterize plaque IPH and LRNC separately, and to identify early sign of plaque micro-calcification which has been recently associated with increased plaque vulnerability [26].

In this study, CTA showed good reproducibility between measurements taken at different time-points. The main causes of variability between repeated measurements were represented by the manual adjustments of plaque contour and vessel lumen, and by the definition between a slightly thickened and a normal vessel. In our analysis the semi-automatic outlining of the outer and inner border of the vessel was reviewed manually to avoid overestimation of plaque LRNC volume that may have been caused by the involuntary inclusion of peri-arterial fat within the volume of interest.

CTA does not have the same limitations as US in the presence of calcium, which has been seen as a plaque feature capable of conferring more stability to the carotid plaque independently from the degree of luminal stenosis [6]. In our analysis we found that a more accurate plaque semi-automatic segmentation is achievable when non-calcified plaques are analysed. Manual adjustment necessary to correct the initial semi-automatic plaque outlining were necessary in 24% of the cases. In these cases, the plaque algorithm applied to the imaging dataset automatically identified both the contrast-enhanced lumen and adjacent plaque calcifications (> 130 HU) as a single volume. To ensure consistency in the CTA plaque analysis and to avoid over/under estimations of calcified plaque volume, we recommend to manually confirm stenosis boundaries on the axial CTA dataset using a window width / window level of 800 / 300 HU, which allows to optimally differentiate the contrast-opacified lumen from plaque calcifications [15]. Therefore, we suggest the use of a dedicated CTA carotid plaque protocol in which the acquisition settings are accurate for both plaque depiction and patient radiation dose safety.

Finally, this study demonstrated that *in-vivo* MDCT assessment of plaque volume and components is feasible with a good intra-reader reliability. Moreover, the results of this study suggest that increased plaque volume may be independently predicted based on the age and gender of the patient. This study showed that larger accumulations of LRNC were found in the moderate stenosis group compared to other NASCET plaque categories suggesting that CTA-HU measurements can facilitate the immediate quantification of key components of carotid atherosclerotic plaque; whose presence, or absence, is linked to plaque vulnerability.

## Data Availability

All data produced in the present study are available upon reasonable request to the authors

## Abbreviations

BIOVASC: Biomarkers Imaging Vulnerable Atherosclerosis in Symptomatic Carotid disease
CPR: Curved-planar reformation
DSA: Digital subtraction angiography
ECST: European Carotid Surgery Trial
FC: Fibrous cap
ICA: Internal carotid artery
ICC: Intraclass correlation
IPH: Intraplaque haemorrhage
LRNC: Lipid rich necrotic core
MRS: Modified Rankin scale
NASCET: North American Symptomatic Carotid Endarterectomy Trial
TIA: Transient ischaemic attack

## Notes

### Competing Interest Statement

The authors have declared no competing interest.

### Funding Statement

This study was partially supported by the Health Research Board (Ireland).

### Author Declarations

This study has been granted full institutional, ethical approval by the Mater Misericordiae University Hospital Ethics Committee on the 2 April 2014 as part of the larger BIOVASC study (Reference number: 1/378/1131). All procedures performed in studies involving human participants were in accordance with the ethical standards of the institutional and/or national research committee and with the 1964 Helsinki declaration and its later amendments or comparable ethical standards.

